# The COVID-19 antibody responses, isotypes and glycosylation: Why SARS-CoV-2 Spike protein complex binding of IgG3 is potentiated in some and immuno-pathologies manifest

**DOI:** 10.1101/2023.01.13.23284524

**Authors:** Raymond Kruse Iles

## Abstract

COVID-19 syndrome does not occur in all who are infected with SARS-CoV-2, and symptoms vary. The anti-SARS CoV-2 Spike immune responses is confounded by the Spike proteins ability to bind Igγ3 heavy chains. This appears to be via sialic acid glycans found on the O-Linked glycosylation moieties of this heavy chain extended neck domain. Furthermore glycosylation of light chains, particularly Kappa (κ), is an associated feature of antibodies binding to SARS-CoV-2 antigens nucleocapsid and Spike protein. COVID-19 recovered patients had increased IgG1 and IgM levels and un-glycosylated κ light chains; possibly In order to counter this immune system subjugation of IgG3. These molecular finding, together with our previous finding that Spike protein binds glycated human serum albumin (HSA), may explain the micro-vascular inflammatory clots that are a causative feature of COVID-19 acute respiratory syndrome (ARDS).

The postulated molecular sequelae are that SARS-CoV-2 virion, entering the blood circulation, being coated with IgG3 and glycated HSA forms a colloid and deposits into micro-focal clots which are also inflammatory. It is not that all IgG3 and albumin is being bound by the virus; this depends on the affinity the SARS-CoV2 virion has for binding an individual’s IgG3 and albumin due to glycosylation and glycation status. The degree of glycosylation and terminal sialyation of an individual’s antibodies is both a genetic and age-maturity dependant feature of the immune system. The degree of HSA glycation is also age related feature particularly related to type 2 diabetes. Thereby establishing the molecular basis of the association of severe COVID-19 disease syndrome and deaths with diabetes, metabolic disorders, and old age. Furthermore, already having cardiovascular disease, with hardened arteries, SARS-CoV2-glycated HSA-IgG3 deposition is going to exacerbate an already compromised circulatory physiology. The binding of IgG3 might also drives a shift in the immune repertoire response to SAR-CoV-2 anti-spike antibodies of increased IgG1 and prolonged IgM levels. This may be associated with Long Covid.

In summary, SARS-CoV-2 Spike protein binding of IgG3, via sialic acid glycan residues, along with increased glycosylated κ-light chains and glycated-HSA may form a focal amyloid-like precipitate within blood vessels which in turn leads to the inflammatory micro-thrombosis characteristic of COVID-19 immuno-pathology.

## 1.0 Introduction

Coronavirus disease 2019 (COVID-19) is an infectious acute respiratory disease (ARDS) caused by a novel coronavirus SARS-CoV2. This virus is airborne, highly contagious and invades respiratory tract tissue. But not all those infected display symptoms and even less (3%) develop severe ARDS [1]. The most common manifestation of severe COVID-19 is pneumonia with fever, cough, dyspnoea and pulmonary infiltrates [1]. Pneumonia can be complicated by respiratory failure requiring oxygen supplementation and mechanical ventilation [2]. Other severe complications include thromboembolism (such as pulmonary embolism and stroke), circulatory shock, myocardial damage, arrhythmias, and encephalopathy [1,3,4]. Severe illness usually develops approximately one week after the onset of symptoms (see figure 1).

**Figure 1.**
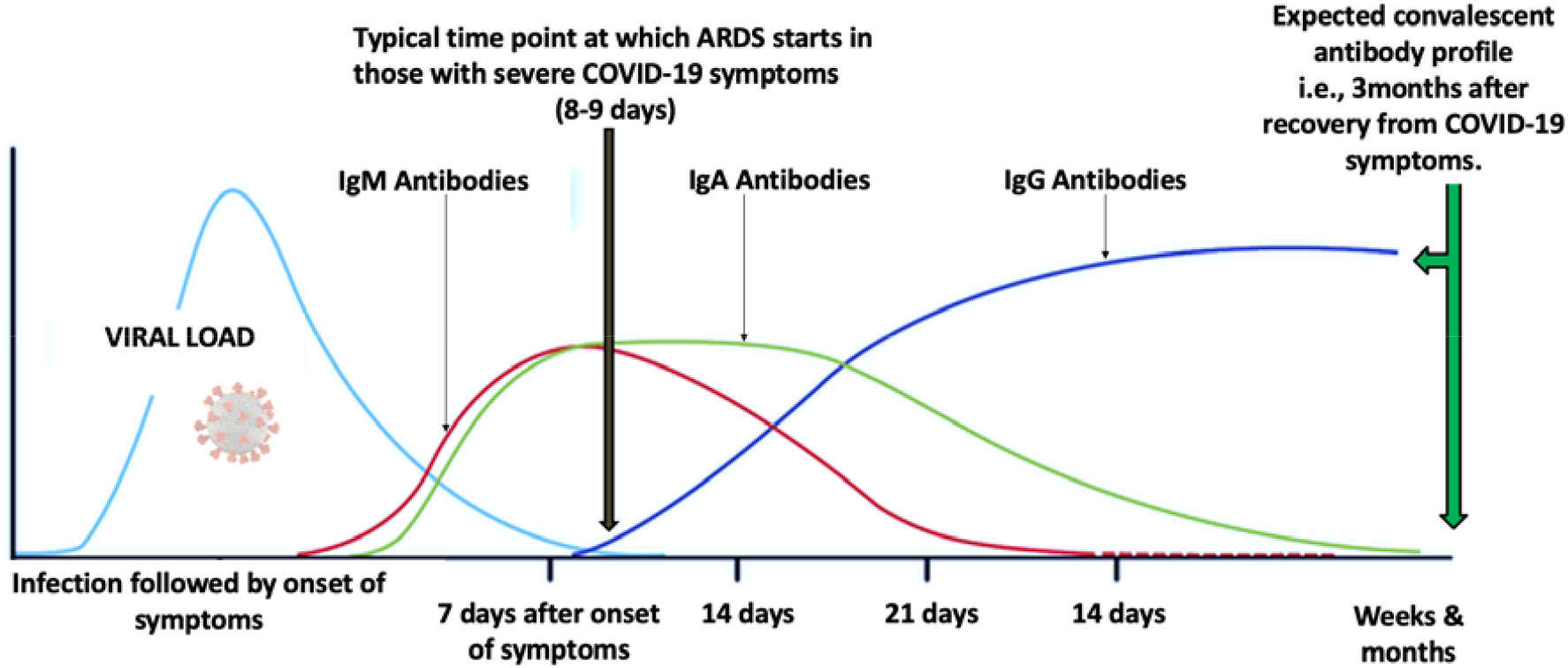
The changing antibody response repertoire after infection with SARS-CoV-2 indicating the time at which ARDS symptoms can appear in COVID-19 and the expected antibody composition profile at convalescence [5-8].

There does not appear to be a lack of antibodies or indeed immune response to SARS-CoV-2 in those who do develop COVID-19 syndrome [5].

### 1.1 The molecular nature of the antibody response

Having circulating antibodies against a pathogen is a gross simplification of the Immunoglobulin biology of the humeral immune response. Aside from the question of what part of the pathogens distinctive molecules, and even then which molecular epitopes, are the responding Immunoglobulin binding; it is important to know what is the isotype and molecular composition of the antibodies being generated? In a basic format antibodies consist of two identical heavy and 2 light chains, joined by disulphide bridges and other non-covalent forces. Given the molecular variant coded by the human genome, this is a combination of heavy chains Igμ, Igα, Igδ, Igε and Igγ and light chain κ and λ. [9] Even amongst the most blood serum dominant of the Immunoglobulin isotypes, IgG; there are four different subtypes of heavy chains (γ1-4), and the secreted mucosal associated Igα heavy chain also exist as two different forms α1 and α2 [10,11]. Furthermore, IgM exists as a pentamer in the blood; whilst IgA when secreted into mucosal fluids exudates, including saliva, is joined to form a dimeric structure [9, 11]. Adding to this Immunoglobulin structural variety, the degree and extent of glycosylation of heavy and light chains increases the molecular complexity of the “antibody response” and associated disease pathologies. Thus, the nature of the antibody response to SARS-CoV2 is a rich, but complex, field of investigation.

However, distinguishing and quantitating the various antibody types arising in a patients serum/ plasma sample to a specific antigen by anti-human immunoglobulin chain assays can be expensive in reagent costs, biological sample volumes required and man hours [12].

Fortunately, the majority of immunoglobulin chains have different molecular masses and we were able to identify and characterise these form a minimal patient sample in a single analysis by highly sensitive MALDI-ToF Mass spectrometry [13,14,15,16,17].

### 1.2 The antibody response in COVID-19

The underlying pathology of COVID-19 ARDS is immunological, with immune suppression agents such as corticosteroids (e.g. prednisolone) showing some benefit [18, 19]. Thus COVID-19 is due to an adverse immune and possibly non-specific response to the SARS-CoV-2 [5,12]. But how and why? The major features of COVID-19 patients appears to be very high levels of anti-SARS-CoV-2 antibodies in the patients who develop COVID-19 syndrome and not a lack of them [12]. In addition the induction of anti-SARS CoV2 antibodies, by Spike protein mRNA vaccinations is effective in reducing incidence of infection and death from SARS-CoV2 in the global population [20,21]. Indeed, *in vitro* analysis of neutralisation have shown that it is having developed anti-Spike antibodies, and not anti-nucleocapsid antibodies, which confer any subsequent protection against reinfection and variants of SARS-CoV-2 [22].

Thus, the contradictory conclusion is that COVID-19 is an immune response pathology characterised by high antibody titres, but induction of anti-SARS spike antibodies by vaccination is protective. The timing at which ARDS symptoms start appears to be when blood serum IgG level to SARS-CoV-2 infection start to rise and IgM antibodies decrease (see Figure 1), but this scenario does presume there has been no prior exposure, the response is completely naive and there is no cross reacting IgG producing B-memory cell activation.

To reconcile this disparity my research team and collaborators, have been looking at the molecular composition of the antibody response in people who did suffer COVID-19 ARDS and those who did not develop COVID-19, despite being at high risk of exposure, or developed only mild symptoms post infection. This was achieved by coupling SARS-CoV-2 nucleocapsid and prefusion conformation Spike proteins to magnetic beads, capturing any bound antibodies and characterising them by MALDI-ToF mass spectrometry (14,15, 17).

### 1.3 COVID-19 Vascular pathologies

There are three major system biology findings of COVID-19 that need to be explained by molecular events. Vascular clots, particular micro clots in pulmonary vessels [23,24], enlarged arteries with echogenic, probably thickened-colloidal, blood flow [25] and a histochemical finding of SARS-CoV-2 virions, or at least Spike protein, can be found at the core of the thrombi [24,26].

In these respects it is important to note Immunoglobulin chain involvement in other vascular diseases. There is an excess production of light chains above, heavy chains by plasma/B-Cells and it is estimated that 40% of these are secreted into the blood circulation as free light chains in inflammatory and autoimmune disorders [27]. This becomes an acute clinical issue in B-Cell malignancies with light chain amyloid deposition in tissues and blood vessels of patients [28]. Studies have shown that both light chains form dimers [29] and also in non-malignant inflammatory conditions such as Multiple Sclerosis [30]. Detailed structural studies have identified a κ-Lc constant domain which is responsible for non-covalent associations in dimerization and other ‘Bence Jones protein’ type oligomerisations [31]. Recent mass spectral based analysis have also shown that glycosylation of light chains is also associated more strongly with myeloma and light chain amyloidosis [32, 33].

We have previous reported our findings concerning the mass spectral analysis of heavy chain and light chain of anti SARS CoV-2 nucleocapsid and Spike antibodies; and also the unusual finding of AGE glycated human serum albumin (HSA) binding by Spike protein [13-17]

Here the complete mass spectral data of light and heavy chain analysis is presented together on the same samples and cross correlated.

Whilst confirmation that IgG3 is a non specific target of prefusion Spike protein binding during viremia is, again, demonstrated; other antibody subtype composition and Immunoglobulin light and heavy chain glycosylation changes, are revealed.

The molecular systems biology of why the biasing of antibody composition & glycosylation, and glycation of serum proteins, in patients infected with SARS-CoV2 leads to COVID-19/ARDS in some individuals, but not the majority, is discussed. Furthermore rapid identification of these features with respect to risk of susceptibility to, and early detection of, COVID-19 syndrome development is proposed.

## 2.0 Material and methods

### 2.1 Samples

Serum and plasma samples were obtained from Health care workers (HCWs) and patients referred to the Royal Papworth Hospital for critical care. COVID-19 patients hospitalised during the first wave and as well as NHS healthcare workers working at the Royal Papworth Hospital in Cambridge, UK served as the exposed HCW cohort (Study approved by Research Ethics Committee Wales, IRAS: 96194 12/WA/0148. Amendment 5). NHS HCW participants from the Royal Papworth Hospital were recruited through staff email over the course of 2 months (20th April 2020-10th June 2020) as part of a prospective study to establish seroprevalence and immune correlates of protective immunity to SARS-CoV-2. Patients were recruited in convalescence either pre-discharge or at the first post-discharge clinical review. All participants provided written, informed consent prior to enrolment in the study. Sera from NHS HCW and patients were collected between July and September 2020, approximately 3 months after they were enrolled in the study.

For cross-sectional comparison, representative convalescent serum and plasma samples from seronegative HCWs, seropositive HCW and convalescent PCR-positive COVID-19 patients were obtained. The serological screening to classify convalescent HCW as positive or negative was done according to the results provided by a CE-validated Luminex assay detecting N-, RBD- and S-specific IgG, a lateral flow diagnostic test (IgG/IgM) and an Electro-chemiluminescence assay (ECLIA) detecting N- and S-specific IgG. Any sample that produced a positive result by any of these assays was classified as positive. Thus, the panel of convalescent plasma samples (3 months post-infection) were grouped in three categories: A) Seronegative HCWs (*n* = 30 samples) B) Seropositive HCWs (*n* = 31 samples); C) COVID-19 Patients (*n* = 38 samples) [34].

### 2.2 Isolation of antibodies and Immunoglobulin analysis

The full process is described in full in our previous publications [14-16]. In summary Recombinant SARS-CoV2 nucleocapsid and recombinant stabilized spike (modified to disable the S1/S2 cleavage site and maintain the pre-fusion stochastic confirmation [13]) were coupled to magnetic beads. Diluted patient serum/plasma was mixed with the respective Nucleocapsid and Spike beads, washed and bound antibodies were eluted at pH2 and disulphide bonds reduced, (5% v/v acetic acid in ddH2O containing 20mM tris(2-carboxyethyl)phosphine (TCEP) - Sigma Aldrich, Poole, UK); thereby liberating the individual antibodies immunoglobulin constituent protein chains for mass spectral analysis.

To minimise problems of individual operator variability, and consistency of process in the target binding proteins recoveries, the Crick automated magnetic rack system was employed. This has been described in full previously [14].

Mass spectra were generated using a 15mg/ml concentration of sinapinic acid (SA) matrix. The eluant from the beads was used to plate with no further processing. The MALDI-ToF mass spectrometer (microflex® LT/SH, Bruker, Coventry, UK) was calibrated using a 2-point calibration of 2mg/ml bovine serum albumin (33,200 m/z and 66,400 m/z) (PierceTM, ThermoFisher Scientific). Mass spectral data were generated in a positive linear mode. The laser power was set at 65% and the spectra was generated at a mass range between 10,000 to 200,000 m/z; pulsed extraction set to 1400ns. A square raster pattern consisting of 15 shots and 500 positions per sample was used to give 7500 total profiles per sample. An average of these profiles was generated for each sample, giving a reliable and accurate representation of the sample across the well. The raw, averaged spectral data was then exported in a text file format to undergo further mathematical analysis.

### 2.3 Spectral data processing and statistical analysis

Mass spectral data generated by the MALDI-ToF instrument were uploaded to an open-source mass spectrometry analysis software mMass™ [35], where it was processed by using; a single cycle, Gaussian smoothing method with a window size of 300 m/z, and baseline correction with applicable precision and relative offset depending on the baseline of each individual spectra. In the software, an automated peak-picking was applied to produce peak list which was then tabulated in Excel. Peak mass and peak intensities tabulated in excel and plotted in graphic comparisons of distributions for each antigen capture and patient sample group. Means and Medians were calculated and, given the asymmetric distributions found, non-parametric statistics were applied, such as Chi^2^ and Kruskal Wallis test, when comparing differences in group distributions.

Peak masses were identified from mass spectra of purified antibodies and human serum albumin as described previously [14,16]. In this study the relationship between immunoglobulin heavy chain molecular mass variants (subtype and glycosylation) was compared to the light chain mass variants for the, each sample and from single sample mass spectra runs. Correlation of intensity of the respective immunoglobulin light and heavy chain was compared with each other per sample and with respect to SARS CoV-2 disease severity grouping.

## 3.0 Results

Of the 99 samples studied 86 generated anti nucleocapsid Ig mass spectra profiles and 88 generated anti spike Ig profiles (see figure 2 panel A).

**Figure 2.**
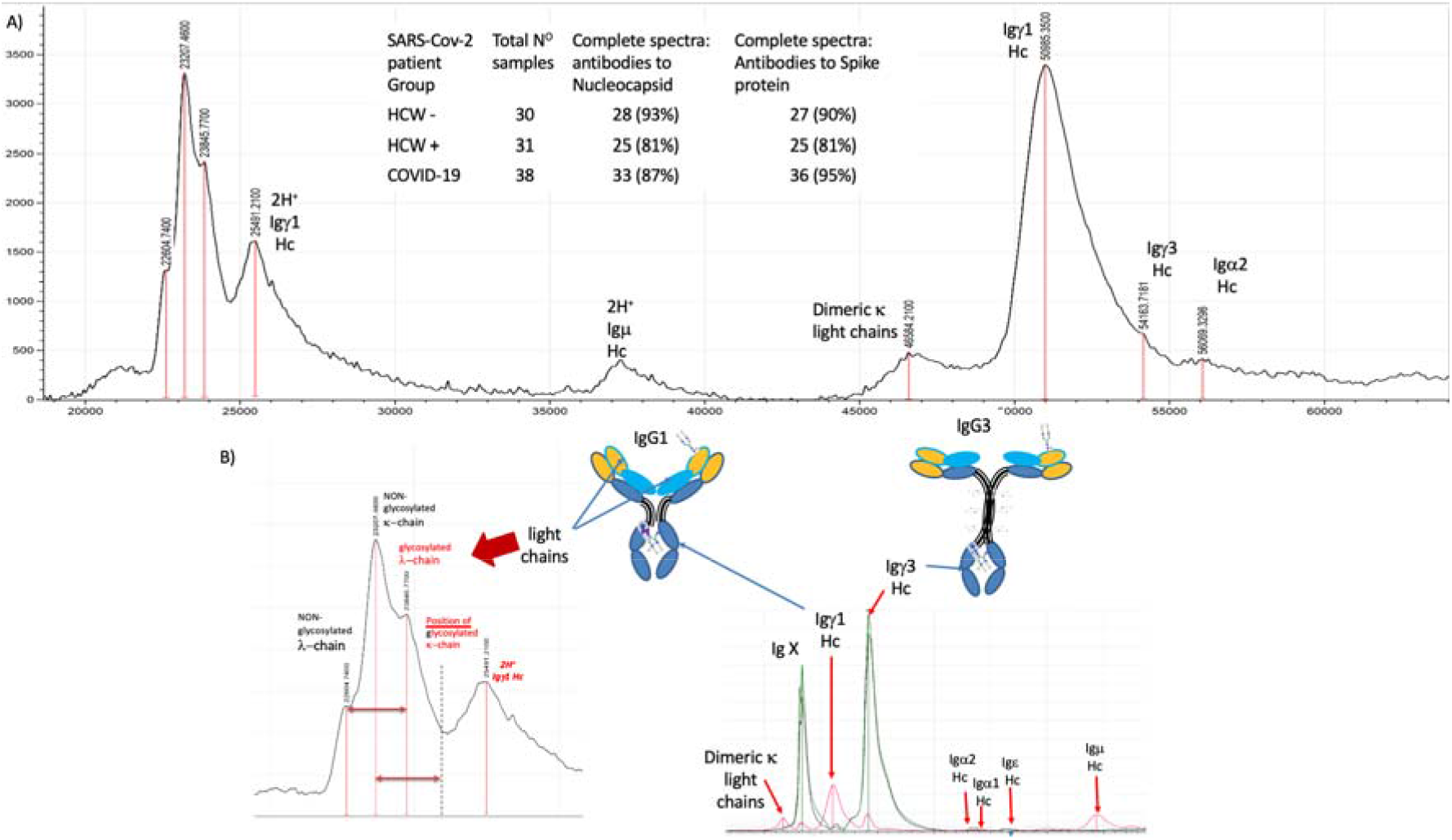
MALDI-TOF mass spectra of anti-SARS CoV-2 antibodies. A) Example spectra of antibody light and heavy chains resolved by MALDI ToF mass spectrometry and proportion of individual samples generating such spectra (inserted table). B) example of how IgG1 and IgG3 light and heavy chains are dissociated by cysteine residue, disulphide bond, reduction and resolved; along with other Immunoglobulins present. The positions of doubly charged and dimeric Ig chains are also indicated.

Following disulphide reduction Ig chains were resolved on the mass spectra as follows, (see example spectra figure 2 panel B). The single charged (H+) immunoglobulin light chains resolved with peak maxima as follows: λ at 22,600-22,800m/z, κ at 23,100-23,300 m/z, glycosylated λ at 23,700 – 24,200m/z and glycosylated κ at 24,800 – 25,200. The heavy chains resolved as Igγ1 at 51,000-51,800 m/z, Igγ3 at 54,000-54,750, Igα1 at 56,100 – 57,500m/z, Igα2 at 55,500-56,100m/z, Igε at 63,400-63,700 and Igμ at 74,200-75,100m/z [13-17].

After deconvolution of doubly charged peak masses, contaminating human serum albumin were identified and had been characterised previously as a higher molecular mass glycated AGE (advanced glycation End products) form, strongly binding to Spike protein. Peaks arising from HSA were not considered in this reported data analysis of immunoglobulins. A novel, ≅ 48,000 m/z, peak found in both nucleocapsid and Spike protein antibody/binding protein preparations was kept within the analysis sets. Not seen on Protein A or G capture of immunoglobulins from the same samples (data not shown), its precise identity is not known. However, this mass equates to a theoretically non-glycosylated Igγ1 heavy chain, and is within 1% mass match of being a theoretical dimeric pair of glycosylated κ and λ light chains of each sample. Dimeric un-glycosylated κ light chain was identified at ≅ 46,000m/z, matching its singly charged form within a 0.2% mass agreement.

### 3.1 Anti-nucleocapsid antibodies

With respect to antibodies to SARS-Cov-2 nucleocapsid; of the light chains, the amount of κ appeared to increases and λ decreased, but this was not statistically significant. With respect to heavy chains Igγ3 was by far the most abundant; detection and levels rose significantly in accordance with severity of symptoms (p<0.01). Much lower than Igγ3 levels, Igγ1 levels did not change between sample groups, Igε levels rose but this increase was not statistically significantly, Igα levels appeared to decrease but again this was not statistically significant. No anti-nucleocapsid Igμ antibodies were detected in these convalescent plasma samples. There was no significant changes in molecular mass of the respective immunoglobulin chains of anti-nucleocapsid antibodies that could be attributed to consistent variation in glycosylation associated with severity of symptoms. (see Table 1 upper panel).

**Table 1.**
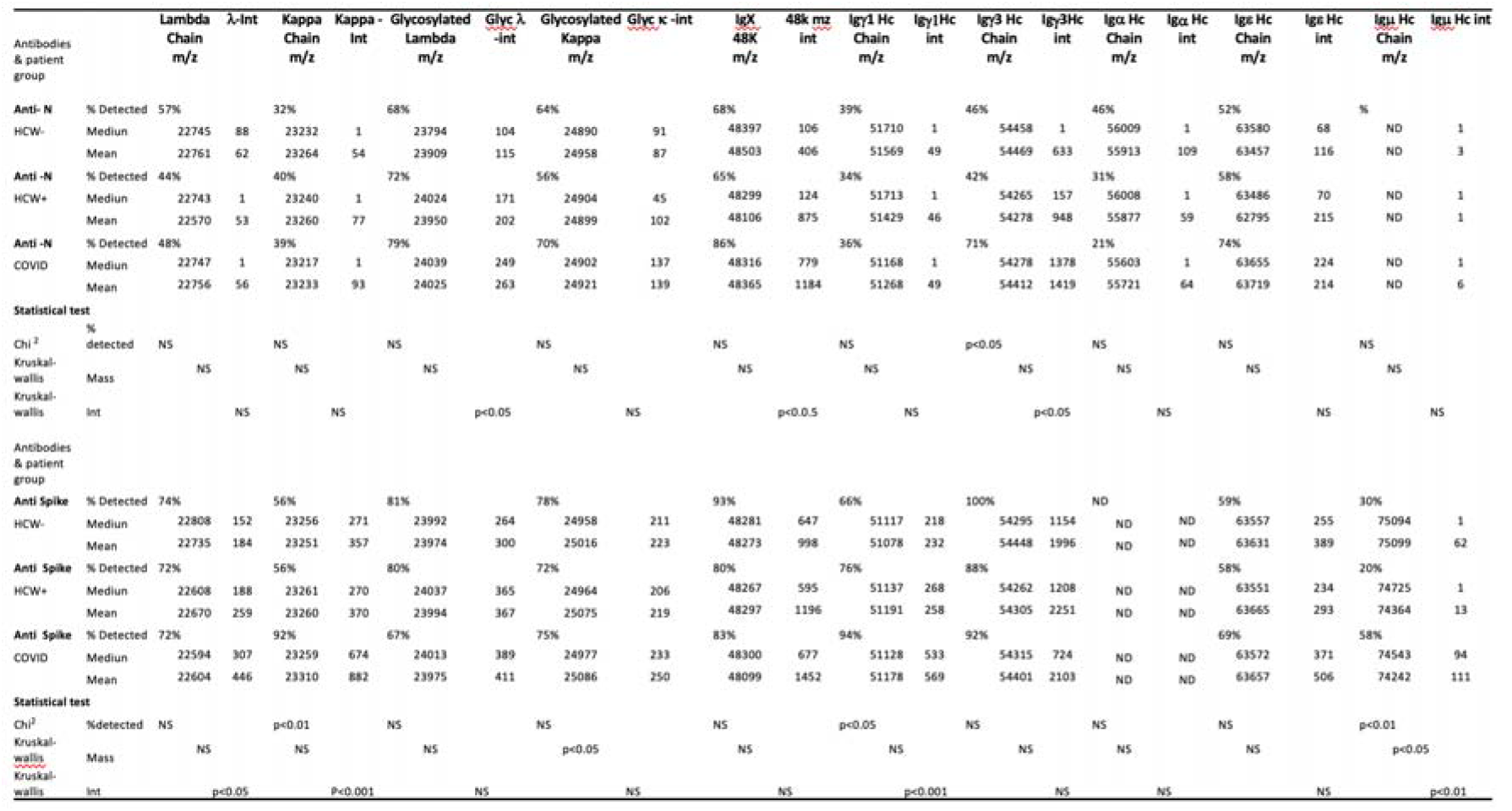
Summary table of anti-SARS-CoV2 immunoglobulin chains detected, (mean and median) mass and intensity for each patient group; along with statistical test comparison significance values for Nucleocapsid antibodies (upper panel) and Spike protein antibodies (lower panel).

Summing the total levels of anti SARS-nucleocapsid light chains detected showed a steady increase with respect to disease severity symptoms (see figure 3a). This was similarly seen for summing of the total immunoglobulin heavy chains (see figure 3a). The increase in total light chains correlated significantly with an increase in total heavy chains (r2= 0.457, figure 3c). However, the heavy chain correlation with disease severity is dominated by the Igγ3 component, which strongly correlated with disease severity (figure 3b) and with total light chain intensity (figure 3b). Indeed, if Igγ3 levels are removed from the heavy chain total the correlation between heavy and light chains are lost (see figure 3c).

**Figure 3.**
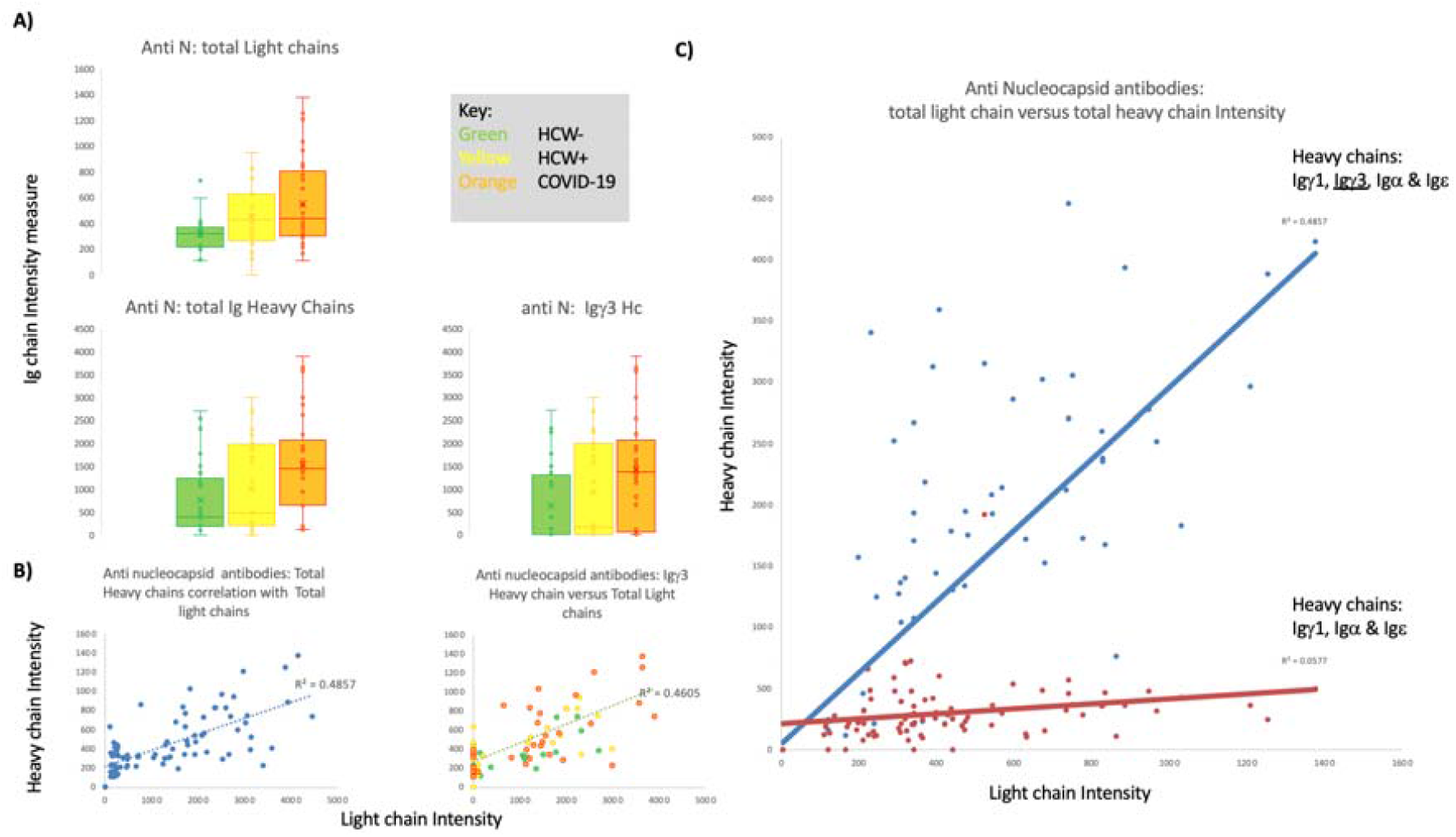
Anti-Nucleocapsid antibodies, total light chain and total heavy chain correlations: Panel A) Box plots of mass spectral intensity levels of total light chains, total heavy chains and Igγ3 for each patient group. Panel B) all patient group correlation plots of total light chains versus total heavy chains (r^2^ =0.4857) and total heavy chains minus Igγ3 levels (r^2^ =0.4605). Panel C) Correlation plots, all patient groups combined, total heavy chains versus total light chains levels; Igγ1, Igγ3, Igα and Igε (r^2^= 0.4857); Igγ1, Igα and Igε (r^2^ =0.0577).

Thus, the matured antibody response to SARS CoV-2 nucleocapsid in convalescent blood plasma/serum is dominated by IgG3. Although the light chains detected in this analytical system could have arisen from any of the reacting immunoglobulins; glycated λ light chains were the most dominant. Similarly the 48,000 m/z peak labelled IgX also showed a statistically significant increases in levels correlating with disease severity (see Table 1 top panel). This peak correlated with both Igγ3 heavy chain levels (see figure 4a) and total light chain levels in all sample groups for the anti-nucleocapsid antibodies (see figure 4b).

**Figure 4.**
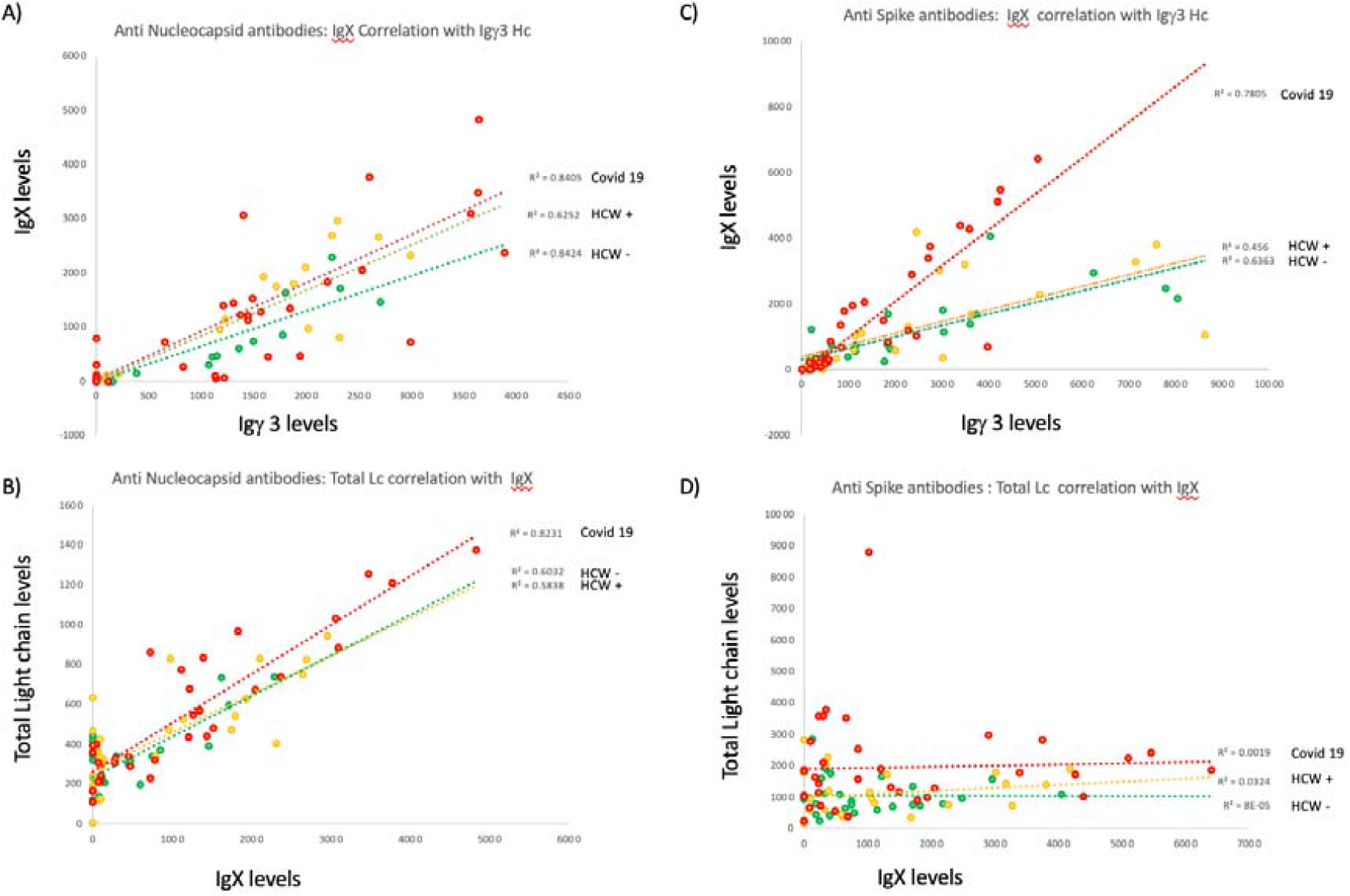
Correlation of 48,000 m/z peak IgX for each HCW-, HCW+ and Covid-19 patient group for Anti-Nucleocapsid antibodies, panel A) and B) and Anti-Spike antibodies, panel C) and D) versus Igγ3 heavy chain (A and C) and total light chains (Band D).

### 3.2 Anti-Spike protein antibodies

With respect to antibodies to the SARS-CoV2 Spike protein, statistically significant increases in both λ (p<0.05) and κ (p<0.01) light chains were seen; the most dramatic increase being in κ light chains (p<0.001) correlating with disease severity. The molecular mass of glycosylated κ light chains showed a small significant increase (p<0.05). However, although Igγ3 was by far the most abundant heavy chain captured it did not show any increase in level correlating with disease severity and was high in all groups. Igγ1, Igε and Igμ, but not Igα, heavy chains were also detected. The levels of Igγ1 and Igμ increased significantly with respect to disease severity (p<0.001 and p<0.01 respectively). In addition Igμ heavy chains showed a small but significant decrease in molecular mass (p<0.01) (See Table1, bottom panel).

When summing the SARS-CoV2 Spike antibody total light chains we see that levels only increase dramatically in the patients recovering from severe COVID-19 ARDS. Summing the total corresponding heavy chains showed no correlation with disease severity and no correlation with total light chain levels (see figure 5a). However, if Igγ3 heavy chain levels are subtracted, the sum of the remaining heavy chain do show an increase but only with respect to COVID-19 ARDS patients (see figure 5b). Furthermore, total heavy chain levels and total light chains strongly correlate in COVID-19 ARDS patients when Igγ3 levels were subtracted (figure 5b). Looking across all samples, including HCW with no reported infection symptoms, only Igγ1 and Igμ heavy chain levels show a significant correlation with total light chain levels (see figure 5c).

**Figure 5.**
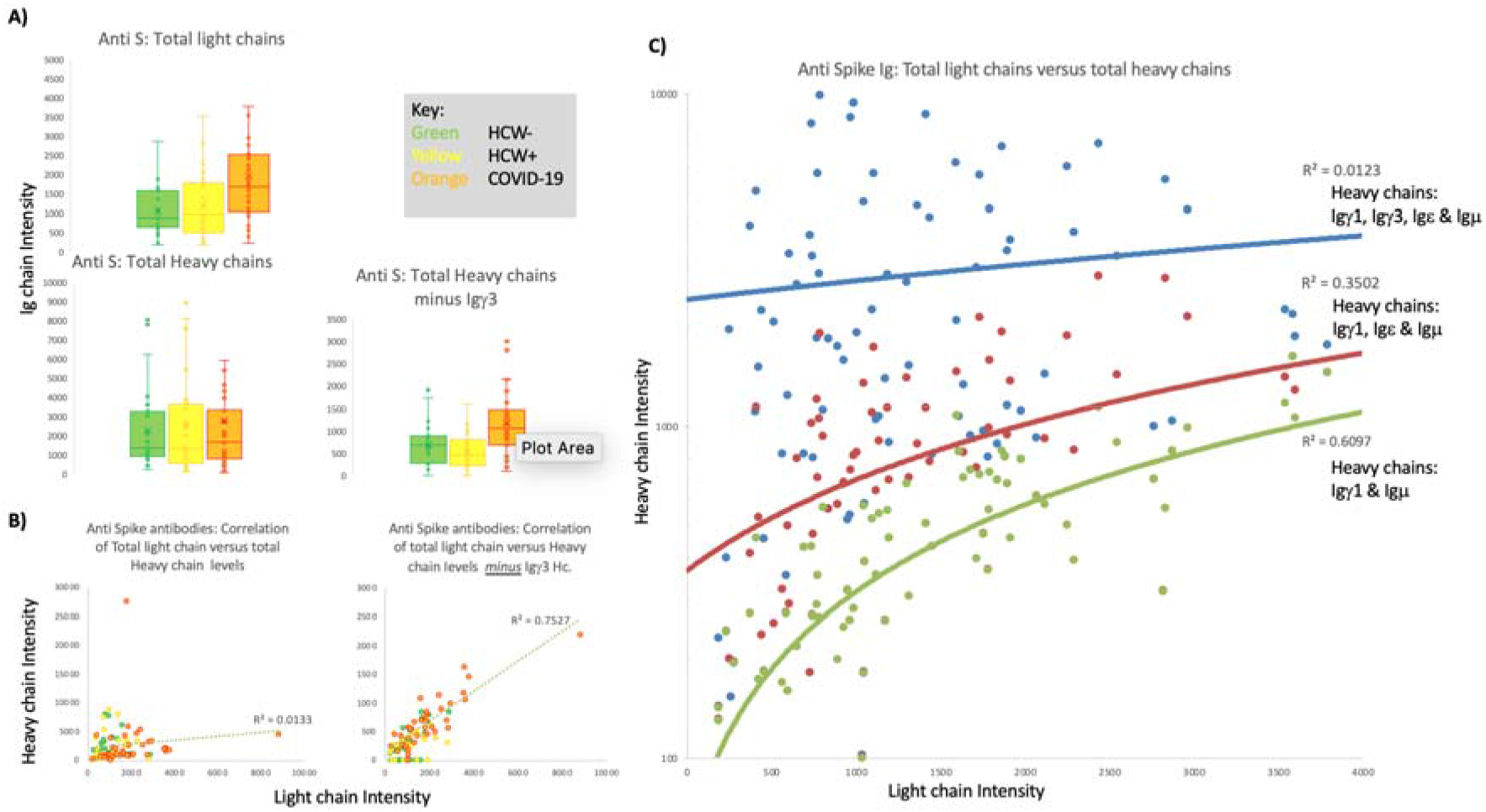
Anti-Spike antibodies, total light chain and total heavy chain correlations: Panel A) Box plots of mass spectral intensity levels of total light chains, total heavy chains and Igγ3 for each patient group. Panel B) Covid-19 patient group correlation plots of total light chains versus total heavy chains (r^2^ =0.0133) and total heavy chains minus Igγ3 levels (r^2^ =0.7527). Panel C) Correlation plots, all patient groups combined, total heavy chains versus total light chains levels; Igγ1, Igγ3, Igε and Igμ (r^2^= 0.0123); Igγ1, Igε and Igμ (r^2^ 0.3502); Igγ1 and Igμ (r^2^=0.6097).

With respect to antibodies against SARS-CoV2 spike protein IgG1 and IgM are the significant response in patients who had developed ARDS and recovered. However, SARS-CoV2 spike protein has a binding affinity for IgG3 this is likely to be via the heavy chain (Igγ3) and we have proposed previously this is due interaction with the extended and O-linked glycosylated neck region [16]. This would appear to be more dominant in patients who develop severe symptoms.

In the convalescent patient samples COVID-19 ARDS patient had to develop elevated levels of Igγ1 and Igμ to deal with the virus. This also appears to be associated with a bias to un-glycosylated light chains in particular Kappa (table 1 lower panel) and a smaller, possibly less glycosylated, Igμ.

For the anti-spike antibodies, the 48,000 m/z peak labelled IgX also showed a statistically significant increases in levels correlating with disease severity (see Table 2, bottom panel). However, this peak correlated with Igγ3 heavy chain levels most strongly in the COVID-19 convalescent patients (see figure 4c) and no longer correlated with total light chain levels in all sample groups (see Figure 4d).

## 4.0 Discussion

The diversity of antibody responses and inflammation in virus infections is a complex web. Antibodies can block key proteins and effectively neutralise their ability to infect a target cell, but also simultaneously recruit other effector immune cells to kill infected cells and alert other tissues to increase their innate intracellular defences via cytokine release i.e. cause inflammation [36]. This is a delicate balancing act with a mixture of antibodies that recruit different inflammatory pathways, along with dampening down certain aspects that do more harm than benefit [37-39].

In this respect, the glycosylation of the antibodies in question has been postulated to modify their pro- or anti-inflammatory ability. This glycosylation can vary on both the light and heavy chains. The consensus is that generally glycosylation of light chains is anti-inflammatory; but hyper glycosylation of the heavy chains is generally pro-inflammatory [40,41,42].

The data presented here strongly indicates a switching to un-glycosylated light chains in patients with Covid ARDS who survived the SARS-CoV2 infection. But this is not the complete story of the antibody differences.

Generation of IgM followed by IgG antibodies is the classic gateway to initiation of an immune response (see figure 1). This is followed by switching to other antibody isotypes and proportional changes to blood-serum antibody isotype compositions. This all is part of a balanced and mature immune response resulting in protective immunity or if appropriate immune tolerance. However, composition biasing and over representation of particular antibody isotypes results in immune pathologies: For example, the over representation of IgE antibodies, reacting to innocuous substances, is a recognised feature of allergies and autoimmunity [43,44].

The development of a mature and memory IgG3 response is common following a viral infection [45,46]. Core nucleocapsids from many viruses are dominant immunogens with high IgG3 titre antibody responses [47,48]. Certainly IgG3 binding to SARS-CoV2 nucleocapsid are likely to have arisen from memory cells previously initiated from prior infection with seasonal human corona virus. Hence the anti-nucleocapsid antibodies detected in a significant number of HCW’s not reporting infection or symptoms. Furthermore, this would appear to also be associated with an increased proportion of glycosylated light chains in the captured antibodies. The unidentified Ig chains labelled IgX and detected at 48K m/z strongly correlated with Igγ3 levels in both anti-nucleocapsid and anti-Spike antibodies, particularly in COVID-19 patient samples. However, the correlation with total light chain levels was only seen in the Nucleocapsid antibodies and not in the Anti-Spike antibodies (see figure 4). This would be consistent with the 48K m/z, IgX peak representing dimers of glycosylated κ & λ light chains; captured as part of non-specific binding of IgG3 by the SARS-CoV-2 Spike protein complex. Since light chain amyloidosis is mostly glycosylated light chains [31-33], and κ-light chain have a far greater propensity than λ to form free circulating light chain dimers [27-30], high levels of free glycosylated κ-light chain dimers are likely to contribute to the COVID-19 ARDS pathology.

Therefore, the dominance of an IgG3 memory response to corona virus nucleocapsid proteins, in particular IgG3 with glycosylated K-light chains, may be detrimental in a SARS-CoV2 infection: The SARS-Spike protein, in its pre-triggered form, binds IgG3. Binding and even cleavage of antibodies is a strategy of many highly dangerous human pathogens to evade and confuse the immune response [49, 50]. Once triggered the tectonic confirmational changes that allows the spike protein to cause enveloped virion fusion with the host cell membrane, would shed the bound IgG3 fragments including light chain dimers [51-53].

Two other important features may make IgG3 antibody binding by SARS-CoV2 more lethal in certain individuals. Binding is strongly implicated to be via terminal sialic glycan residues. The degree and exact sialic acid linkage is controlled by a number of different enzymes whose expression is controlled by genetic factors and age [54-57]. In addition metabolic disorders such as diabetes result in non-enzymatic glycation of serum proteins resulting in sialic like positive charges being deposited on blood serum proteins [58]; the most abundant being albumin followed by Immunoglobulins [59].

In this report we first show that COVID ARDS patients have a high level of bias toward IgG3 antibodies in their response to SARS-CoV2 nucleocapsid antigen. Second we have demonstrated SARS -CoV2 Spike binding significant amounts of Igγ3 and a bias towards high levels of glycosylated light chains, associated with that Igγ3 heavy chain. Third that COVID-19 ARDS patients who have recovered, had increased levels of Igγ1 and Igμ associated with increased un-glycosylated Kappa light chains in their convalescent blood serum/plasma antibody response to SARS-CoV2 Spike protein.

Although IgG1 and IgM may represent a more naive inflammatory response to the SARS CoV-2 Spike protein complex, this is, in all probability, a recovery consequence in the COVID-19 ARDS clinical phenotype. The pathology arising in some individuals being the sequalae of SARS-CoV2 Spike protein binding of IgG3, particularly hyperglycosylated IgG3, and glycated albumin. These bound IgG3 antibodies with the virus, or if shed when the spike complex is triggered by ACE-2 receptor binding, will precipitate [60,61,62]. Together this complex of Igγ3, glycosylated κ light chains and glycated albumin would be released and deposited within the vasculature [63,64]. This would not only represent a source-site for complement fixation & exhaustion [65] but also for clots [66] and even dilation; with interstitial tissue fluid movement into vessels along with proteinuria [67 - 70]. In a previous report we demonstrated, with the same samples, increased glycated human serum albumin is a feature of the serum/plasma of these same COVID-19 ARDS convalescent patients [14].

## 5.0 Conclusion

IgG3 is a major feature in the biology of SARS-CoV2 patho-physiology. It is the dominant blood serum antibody produced in response to SARS nucleocapsid, but is bound by SARS-CoV-2 spike complex. This is likely to be via increased sialic acid residue terminal glycation of O-linked glycans found on the extended neck domain of Igγ3. Thus, individuals with a genetic/epi-genetic predisposition to increased terminal sialic acid glycosylation will have IgG3 bound more strongly. If combined with glycated albumin and increased glycosylated Kappa light chains precipitation within the vasculature, the resultant complex will be a focus for inflammation and clots [23, 24, 66] (see figure 6).

**Figure 6.**
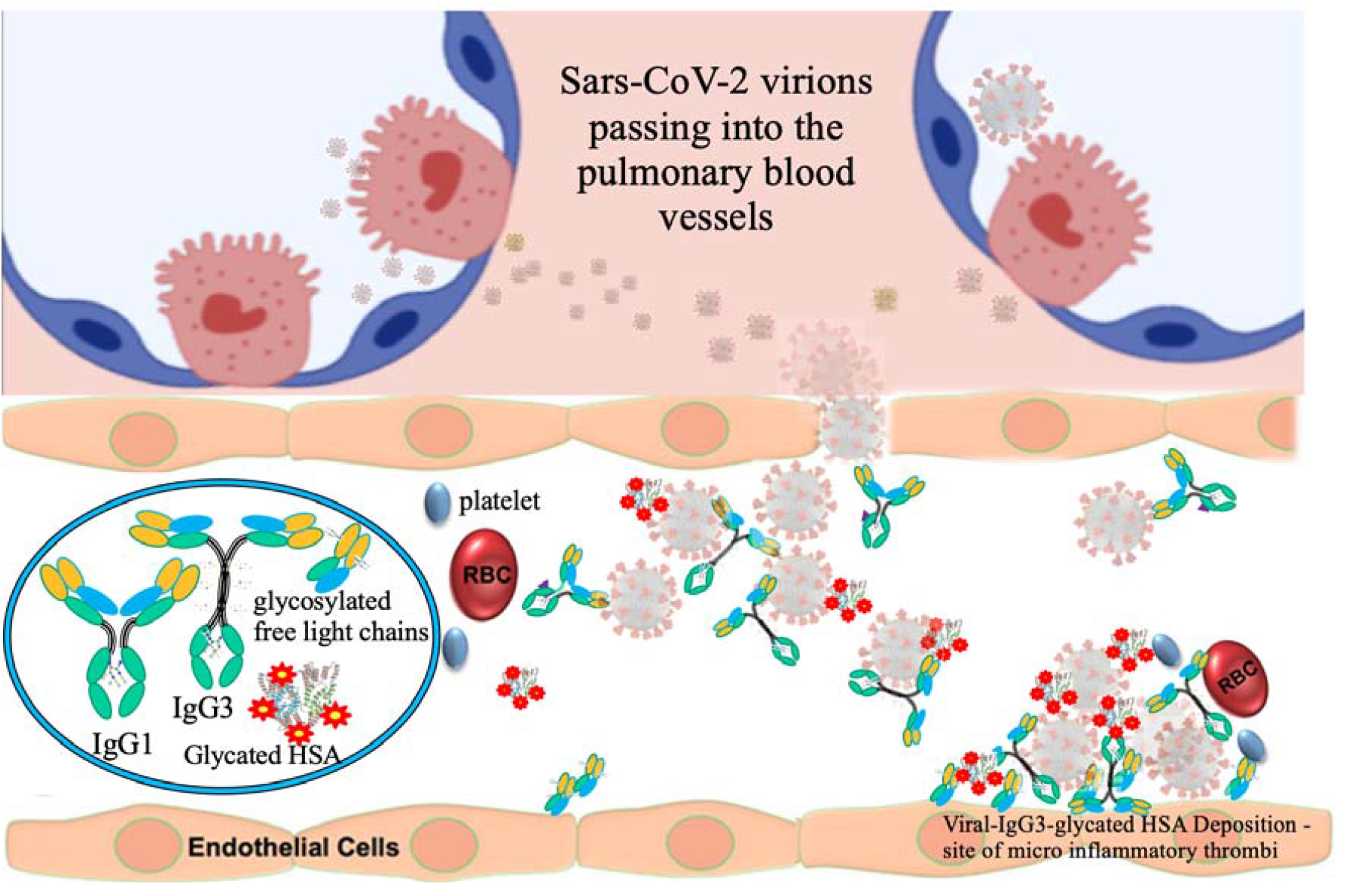
Diagrammatic illustration of a proposed mechanism by which SARS-CoV-2 virion pass into pulmonary blood vessels from infected alveoli where they encounter glycated albumin and IgG3 which are bound non-specifically; along with specific binding of IgG1 to the viral envelope Spike glycoproteins. These form a colloidal clumps of virion-IgG3-glycatedHSA and precipitate forming thrombotic-inflammatory clots along with enmeshed platelets and RBC. This may be assisted by vascular foci deposited glycosylated free (dimeric) light chains arising from high antibody titres.

With IgG3 inactivation those destined to develop COVID-19 symptoms switch back to a more naive IgG1 and IgM antibody response in order to combat SARS-CoV2 viremia. Measuring the precise molecular glyco-composition of key anti-SARS immunoglobulins in patients’ blood, along with glycated albumin levels, may strongly identify those most at risk of developing COVID-19 ARDS.

## Data Availability

All data produced in the present study are available upon reasonable request to the authors

## Abbreviations

HCW: Health care workers
ARDS: Acute respiratory distress syndrome
ITU: Intensive Therapy unit
AGE: advanced glycation end products
MALDI-ToF: Matrix assisted laser desorption ionization – time of flight
Ig: immunoglobulin protein chain

## Acknowledgements

Thanks go to the Humoral Immune Correlates to COVID-19 (HICC) consortium, funded by the UKRI and NIHR; grant number G107217 (COV0170 - HICC: Humoral Immune Correlates for COVID-19) for providing samples. To Jason Iles, Raminta Zmuidinaite, Jonathan Lacey and Anna Gardner for generating the mass spectral analysis of samples as detailed in prior publications.

I thank the Royal Papworth Hospital Foundation Trust COVID-19 Research and Clinical teams led by Dr Helen Baxendale and including Intensive care clinicians, Catherine Denny, Maria Nizami, Erin Hopley for supporting recruitment of patients and HCWs who participated.

## Author Contributions

Conceptualisation, formal data analysis, investigation, data curation, writing—original draft preparation, writing—review and editing, visualisation, R.K.I. The author has read and agreed to the published version of the manuscript.

## Institutional Review Board Statement

The study sample collection and overall aims was approved by the NHS Research Ethics Committee, Wales, IRAS: 96194 12/WA/0148. Amendment 5.

## Informed Consent Statement

NHS HCWs participants from the Royal Papworth Hospital were recruited through staff email as part of a prospective study to establish seroprevalence and immune correlates of protective immunity to SARS-CoV-2. Patients in convalescence were recruited either pre-discharge or at the first post-discharge clinical review. All participants provided written informed consent prior to enrolment in the study.

## Data Availability Statement

Compiled summary data can be made available upon request to the corresponding author. Raw mass spectral data from the individual samples will require compiling from archives at MAP Sciences and so require a detailed project proposal to justify the additional resource expenditure required in providing this complete data set.

